# Cosmic signature SBS39 is associated with homologous recombination deficiency

**DOI:** 10.1101/2024.10.23.24316019

**Authors:** Yuan Chun Ding, Shu Tao, Allen Mao, Elad Ziv, Susan L Neuhausen

## Abstract

Cosmic single-base-substitution mutational signature 3 (SBS3) is associated with hereditary and somatic mutations in genes involved in homologous recombination deficiency (HRD) and predicts response to PARPi. The COSMIC database was updated from 30 SBS signatures in version 2.0 (V2.0) to 86 SBS signatures in Version 3.4 (V3.4). We found that SBS3 in V3.4 was poorly associated and SBS39 was strongly associated with germline and somatic mutations in HRD genes and should be classified as an HRD signature.

## REPORT

Six different classes of mutational signatures have been cataloged in the database of Cosmic mutational signatures[1, 2]. Single base substitution signature 3 (SBS3) in Cosmic signature version 2 (V2.0) is associated with homologous repair deficiency (HRD) and the majority of individuals with pathogenic germline and somatic *BRCA1/BRCA2/PALB2* mutations have this signature[3-5]. Tumors with pathogenic mutations in these genes are known to impair repair of double-stranded DNA breaks (DSBs) and have increased sensitivity to treatment with poly-ADP-ribose (PARP) inhibitors[6]. Therefore, identification of tumor samples with SBS3 is clinically important for targeted treatment.

Recently, the COSMIC SBS signatures were expanded to 86 signatures in Version 3.4 (V3.4) from 30 in V2.0 (https://cancer.sanger.ac.uk/signatures/sbs/). In an analysis of signatures from breast tumors, we observed that SBS3 in V3.4 was much less frequent than SBS3 in V2. Therefore, we compared V2 and V3.4 and investigated whether there were other signatures in V3.4 that were associated with HRD by capitalizing on the strong association of the HRD signature and pathogenic germline and somatic mutations in *BRCA1/BRCA2*/*PALB2*.

We used data from whole exome sequencing of 763 tumor/normal pairs of female breast cancer patients seen at the City of Hope Medical Center (detail in on-line methods) and data from the ovarian serous cystadenocarcinoma data from The Cancer Genome Atlas (TCGA)[7] downloaded from the cbioportal (https://www.cbioportal.org/) database. We assigned SBS signatures for V2.0 and V3.4 using two different methods, MutationalPatterns[8, 9] and SigProfilerAssignment[10] (details in on-line methods).

Individual mutational signatures for each version are provided in Supplementary Table 1. Of the 763 breast cancer patients, 43 carried pathogenic germline mutations and an additional 15 patients had truncating somatic mutations in *BRCA1, BRCA2*, and *PALB2*. We first selected signatures in V3.4 relevant to those germline and somatic mutations using a random forest classification and feature selection algorithm (see On-line Methods), implemented in the Boruta R package[11]. For signatures assigned by the two methods, SBS39, SBS8, and SBS3 were the top signatures positively associated with *BRCA1*/*BRCA2*/*PALB2* mutations and SBS39 had a higher rank of importance than SBS3 (Supplementary Figure 1). The area under the ROC curve (AUC), optimal cut point by the Youden index, and results of Fisher’s exact test of association between *BRCA1*/*BRCA2*/*PALB2* mutation status (Yes or No) and signature value (Low or High) dichotomized by the optimal cut point for each signature are in Table1 and Supplementary Table 2. For V3.4, SBS39 was the only signature with a relatively high AUC that was positively associated with mutation status (AUC = 0.81 and 0.64 for MutationalPatterns and SigProfiler, respectively) (Supplementary Table 2); SBS3 and SBS8 also showed some association with mutation status but their AUC values were much lower than the AUC value of SBS39 (Table 1, Fig. 1b-1c, and Supplemental Table 2). In V2.0, only SBS3 showed high AUC values (AUC = 0.8 and 0.85 by SigProfiler and MutationalPatterns, respectively) (Table 1 and Supplementary Table 2). Compared to SBS3 V2.0, SBS3 V3.4 showed much weaker association with mutation status (Wilcoxon test) (Supplemental Figure 2a-d). In the SigProfiler signature assignment, 143 of 763 samples had SBS3 in V2.0, but only 22 of those 143 samples had SBS3 in V3.4; of those 121 samples without SBS3, 50 had SBS39. Eight of 58 samples with SBS39 V3.4 did not have SBS3 in V2. About half (72 of 143) of SBS3 V2.0 signatures had split into SBS3 (22 of 143) and SBS39 (50 of 143) in V3.4. Interestingly, for signature V3.4 values assigned by SigProfiler, there was mutual exclusivity of SBS3, SBS8, and SBS39 for all but two samples (middle panel of Figure1a). This pattern of mutual exclusivity allowed us to create new signature variables SBS39n3 and SBS39n3n8 by the sum of the two or three corresponding signature values for all but the two samples without mutual exclusivity. These combined signatures had higher AUCs compared to individual signatures with the AUC increasing to 0.71 from 0.64 in SBS39 (Table 1 and Figure 1b). The strictly exclusive signature patterns were not observed in signatures assigned by MutationalPatterns although SBS3V2.0 was not observed for most of samples in the expanded V3.4 SBS signatures (Supplementary Table 1). The significance of association between SBS39n3n8V3.4 assigned by SigProfiler and mutation status was similar to that between SBS39V3.4 assigned by MutationalPatterns and mutation status (Fig.1d-e). Compared to the no-mutation sample group, both signatures showed significantly higher signature values in the germline-mutation sample and truncating-somatic-mutation sample groups, with non-significant differences between those two mutation groups. (Fig.1d-e).

**Table 1.**
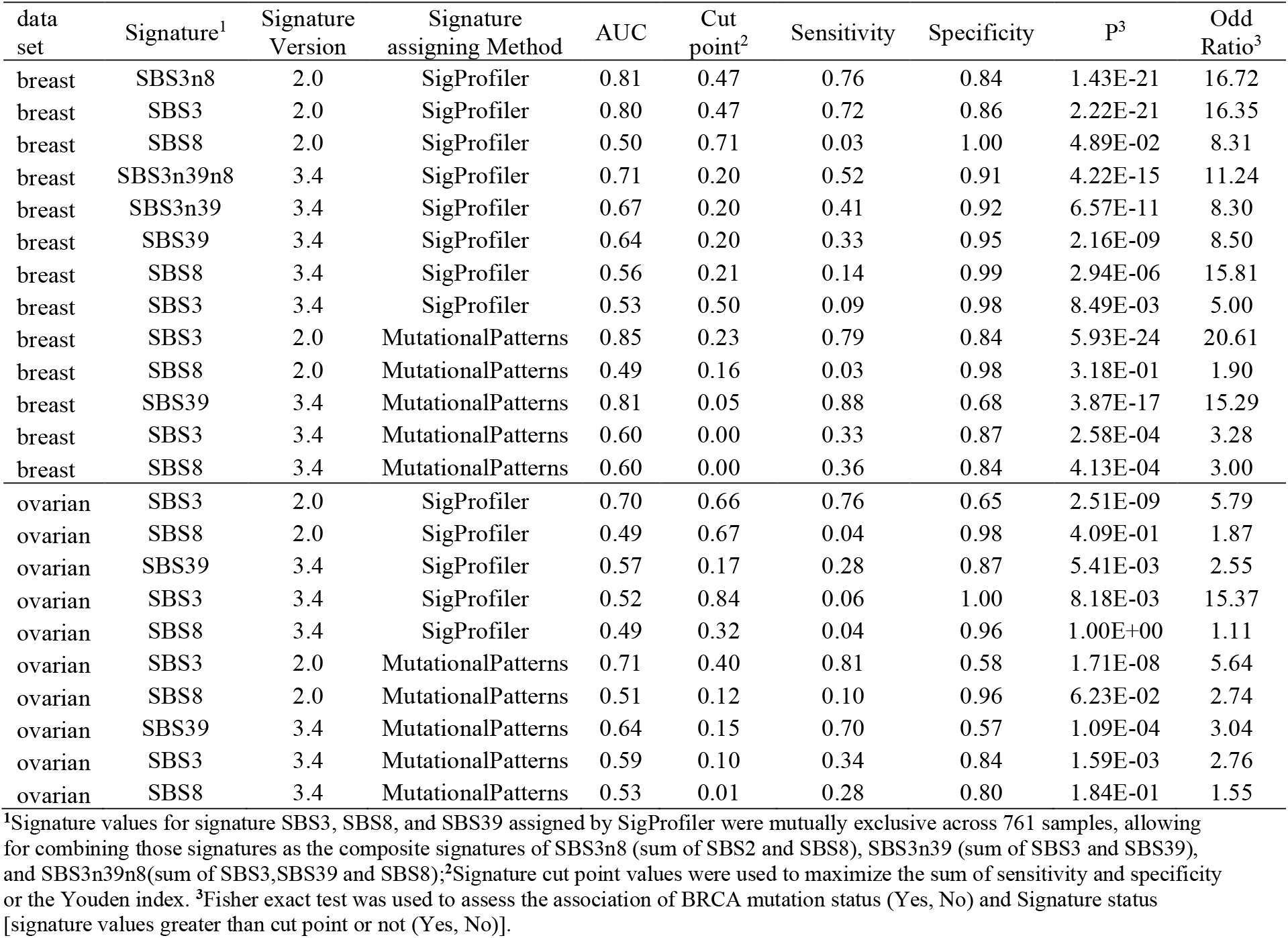
Association of BRCA mutation status and presence of Signature 3, 8 and 39 in two data sets.

**Fig. 1.**
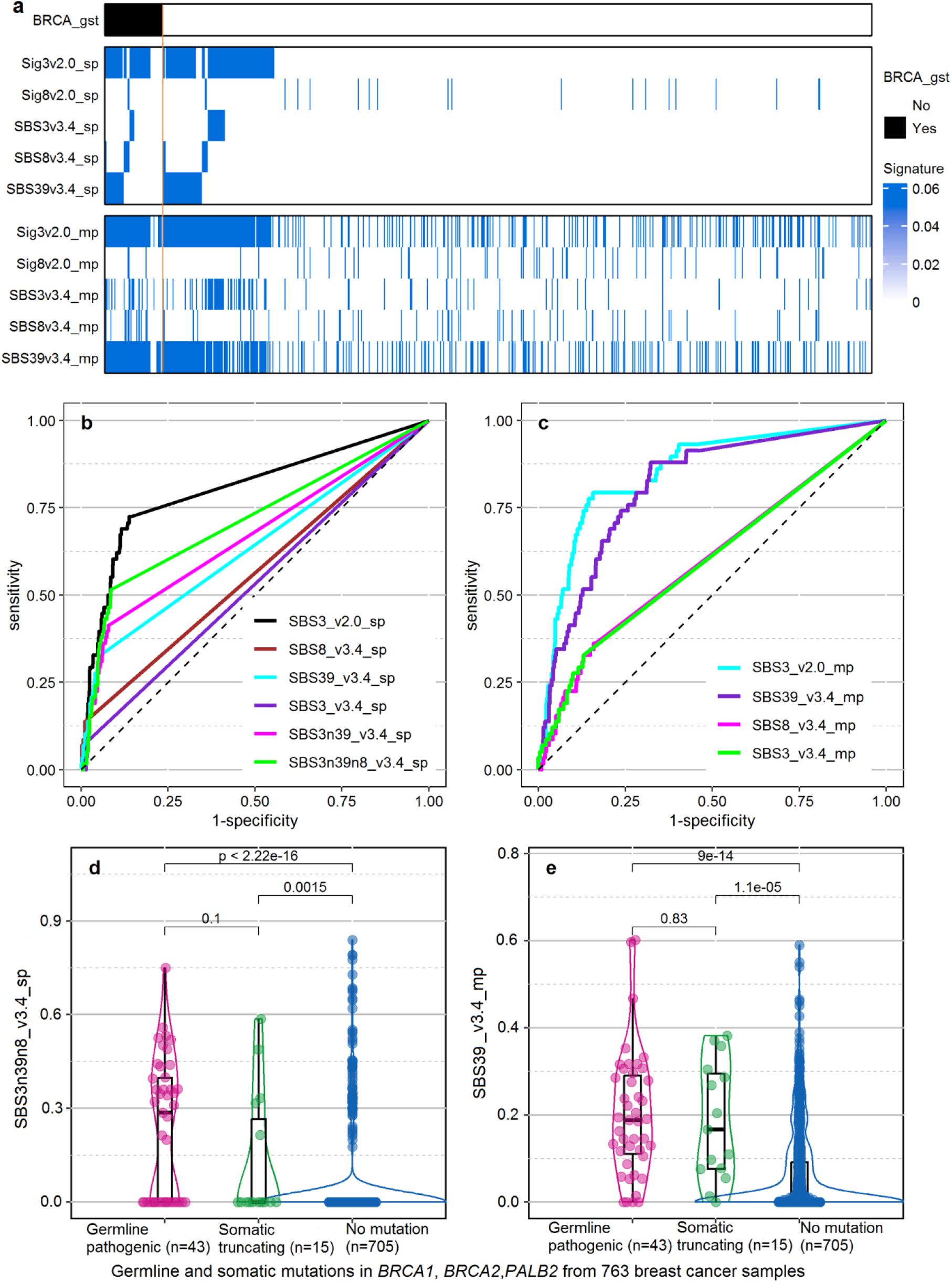
SBS39 is associated with pathogenic germline and truncating somatic mutations in *BRCA1/BRCA2/PALB2* from 763 breast cancer samples. **a**. Heatmap of pathogenic germline and truncating somatic mutations (BRCA_gst) (top); single-base-pair mutational signature 3 V2.0 (SBS3 v2.0), signature 8 V2.0 (SBS8v2.0), SBS3 version 3.4 (SBS3v3.4), SBS8 version 3.4 (SBS8v3.4), SBS39 version 3.4 (SBS39v3.4) assigned by SigProfiler (sp) (middle) and MutationalPatterns (mp) (bottom). **b**. the receiver operative characteristic (ROC) curves for SBS3v2.0 and SBS3, SBS8, SBS39 of v3.4 and the combination of SBS3 and SBS39 (SBS3n39), and SBS8 (SBS3n39n8) assigned by SigProfiler. **c**. ROC curves for SBS3v2.0 and SBS3, SBS8, SBS39 of v3.4 assigned by the MutationalPatterns. **d**. Scatter-box-violin plots of relative signature values, assigned by SigProfiler, for the combined signatures (SBS3n39n8v3.4) stratified by mutational status of *BRCA1/BRCA2/PALB2*; association of signature with mutation status was tested by the Wilcoxon test. **e**, Scatter-box-violin plots of SBS39v3.4, assigned by MutationalPatterns and its association with mutation group status (the Wilcoxon test).

For the TCGA ovarian cancer data [14], 50 of 313 samples had pathogenic germline mutations and 17 additional samples had pathogenic somatic mutations (Supplementary Table 4 and On-line Methods). We found similar association patterns between signature and mutation where the highest AUC was for signature SBS3V2.0 and in V3.4, SBS39 was the new signature associated with *BRCA1/BRCA2/PALB2* mutation status whereas there were many fewer samples with SBS3 and a much weaker association with mutation status. AUC values were lower in the ovarian data than the breast data (Supplementary Table 5 and Table 1).

Diaz-Gay M. et al tested performance of signature assignment of five tools for signature refitting using the complete set of 79 signatures in V3.3 as input [10]. They found that SigProfiler showed the highest precision with relatively high sensitivity and the standard mode of MutationalPatterns had the highest sensitivity and the worst precision among the five tools[10]. Low precision generally is due to signature overfitting, which can be improved by only refitting a subset of signatures that is specific to a target data set[12]. For signature assignment using the MutationalPatterns, we refitted a subset of signature specific to the breast or ovarian cancer data (see Online methods). In general, higher AUC values, higher sensitivity, and lower specificity values were observed from MutationalPatterns compared to SigProfiler (Table 1, Supplementary Tables 2 and 5). In the context of use of mutational signatures for selection of patients for PARP inhibitor treatment, one may want increased sensitivity even if there is lower specificity because response to treatment is observed in patients without *BRCA1/BRCA2/PALB2* mutations[13]. We focused on the association between SBS signatures in V3.4 and pathogenic mutations in *BRCA1, BRCA1*, and *PALB2*, genes known to be associated with HRD. Prior studies have demonstrated that having pathogenic mutations in these genes also are good predictors of response to PARPi[14, 15]. Although other mechanisms (epigenetic silencing, somatic mutation and/or copy number loss) and other genes also may drive tumors with HRD, this would likely diminish the specificity but not the sensitivity of our results. Therefore, comparison of sensitivities using the different signature profiles will likely represent a reasonable approximation of response to PARPi.

In this study, we observed that SBS3V2.0 was most closely correlated with SBS39 in V3.4 in the expansion of the number of signatures from V2.0 to V3.4 with some residual correlation with SBS3 and SBS8. This potentially may be explained by the mutation profile of the six mutation types for each signature as shown in Supplementary Figure 3. For example, SBS39V3.4 and SBS8V3.4 were characterized by the specific contribution of C>G and C>A mutations, respectively whereas, SBS3V2.0 was characterized by the inclusion of both mutation types. Regardless of the reason, when using Cosmic SBS signatures V3.4 for determining HRD status, SBS39 is the most informative signature for HRD and SBS39, SBS8, and SBS3 all should be included to optimize prediction of HRD. SBS39 should be classified as an HRD signature rather than a signature with unknown etiology.

Further replication of its association with HRD can be done using additional exome and whole genome sequencing data for breast and ovarian cancer samples. Further work, ideally in the setting of patients treated with PARPi, is required to optimize the SBS signatures and/or combine with other mutational processes to predict response to PARPi.

**Supplementary Figure 1.**
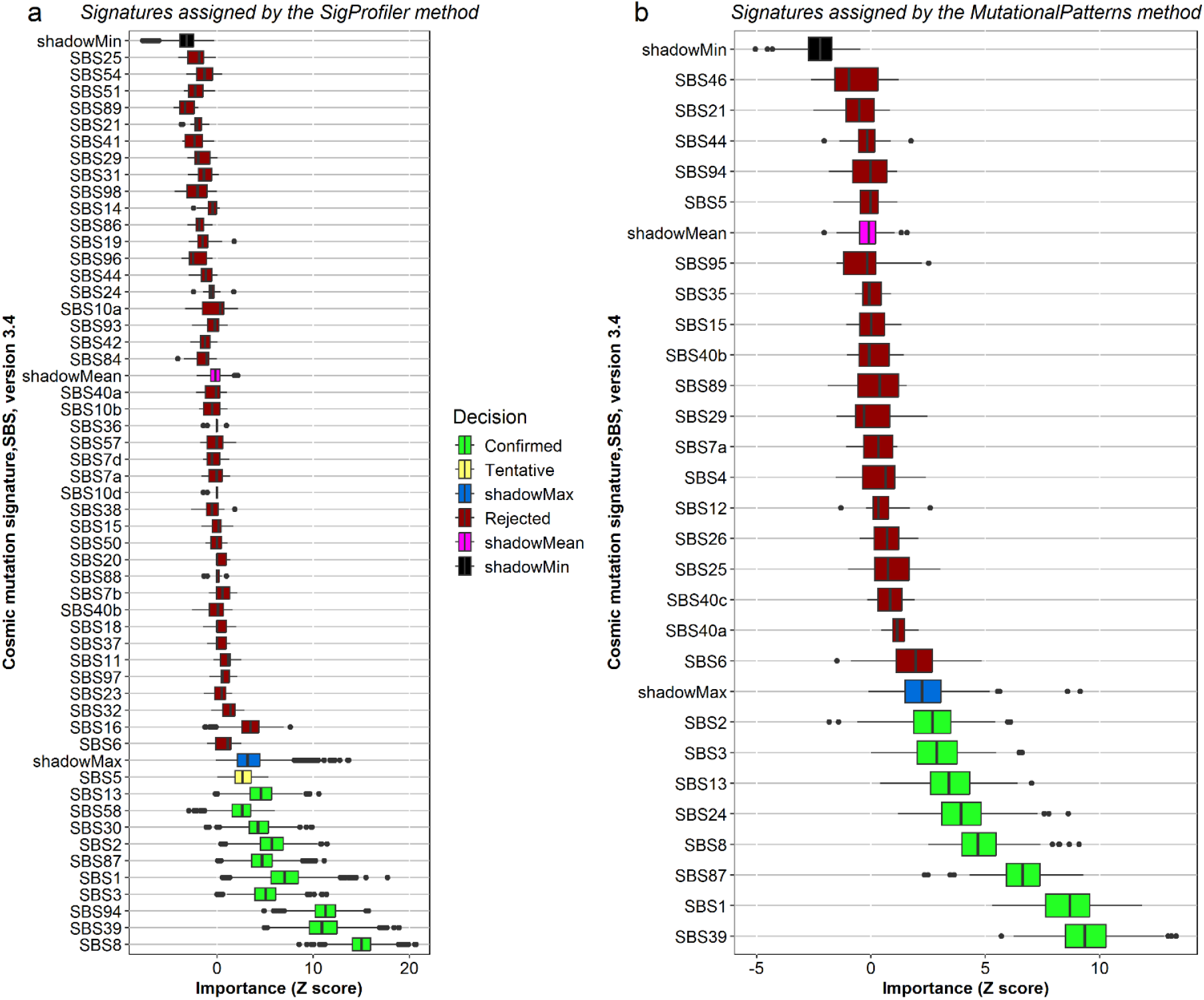
Selection of version-3.4 signatures relevant to *BRCA1*/*BRCA2*/*PALB2* mutations. A random forest classification and feature selection algorithm, implemented in the Boruta R package, was used to select signatures relevant to *BRCA1*/*BRCA2*/*PALB2* mutations for version 3.4 signatures from 763 breast cancer samples. Signature relevance to mutation was determined by comparing relevance of real signature to that of its shadow signature created by randomly shuffling values of the original signature across samples (see On-line Methods). Plot **a** is for signatures assigned using the SigProfile method; plot **b** for signatures assigned by the MutationalPattern methods. Signatures represented by green boxplots and the Z score (relevance measure) on the x axis greater than the maximal relevance score of shadow signatures (see the “shadowMax” in the decision legend in the middle of two plots) were considered as signatures relevant to *BRCA1*/*BRCA2*/*PALB2* mutations. SBS1 and SBS87 were negatively associated with *BRCA1*/*BRCA2*/*PALB2* mutation since higher signature values were enriched in samples free of mutation; the rest of signatures with the green box showed positive associations. SBS5 represented by the yellow boxplot in plot a was a tentative important signature relevance to mutation among the signature assigned by the SigProfilerAssignment tool.

**Supplementary Figure 2.**
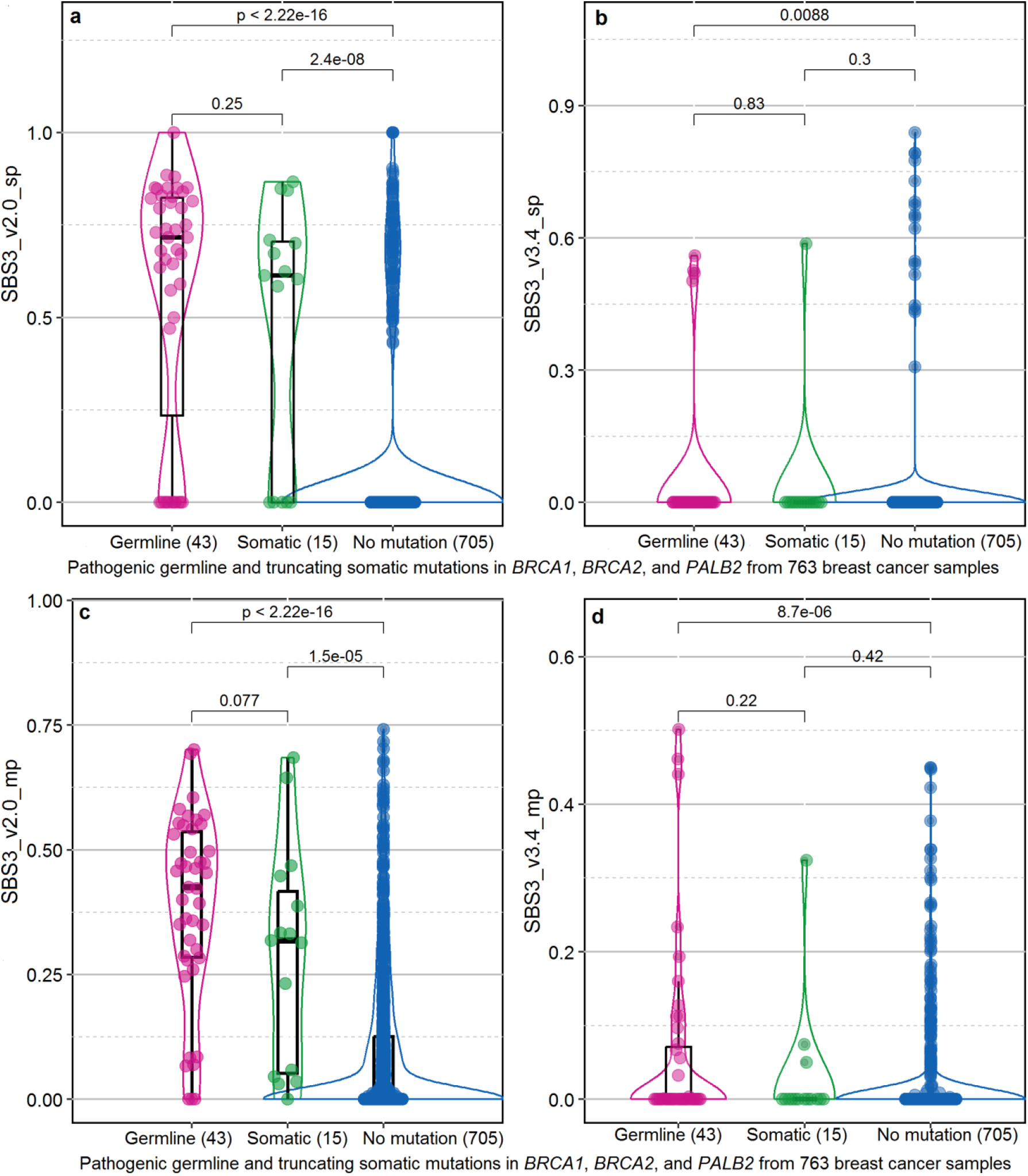
Compared to SBS3 V2.0, SBS3 3.4 showed much weaker association with *BRCA1/BRCA2/PALB2* mutations (Wilcoxon test). **a**. Scatter-box-violin plots of SBS3 V2.0 assigned by SigProfiler. **b**. Scatter-box-violin plots of SBS3 V3.4 assigned by SigProfiler. **c**. Scatter-box-violin plots of SBS3 V2.0 assigned by MutationalPatterns. **d**. Scatter-box-violin plots of SBS3 V3.4 assigned by MutationalPatterns. the Wilcoxon test was used to test association of signature with mutation group status.

**Supplementary Figure 3.**
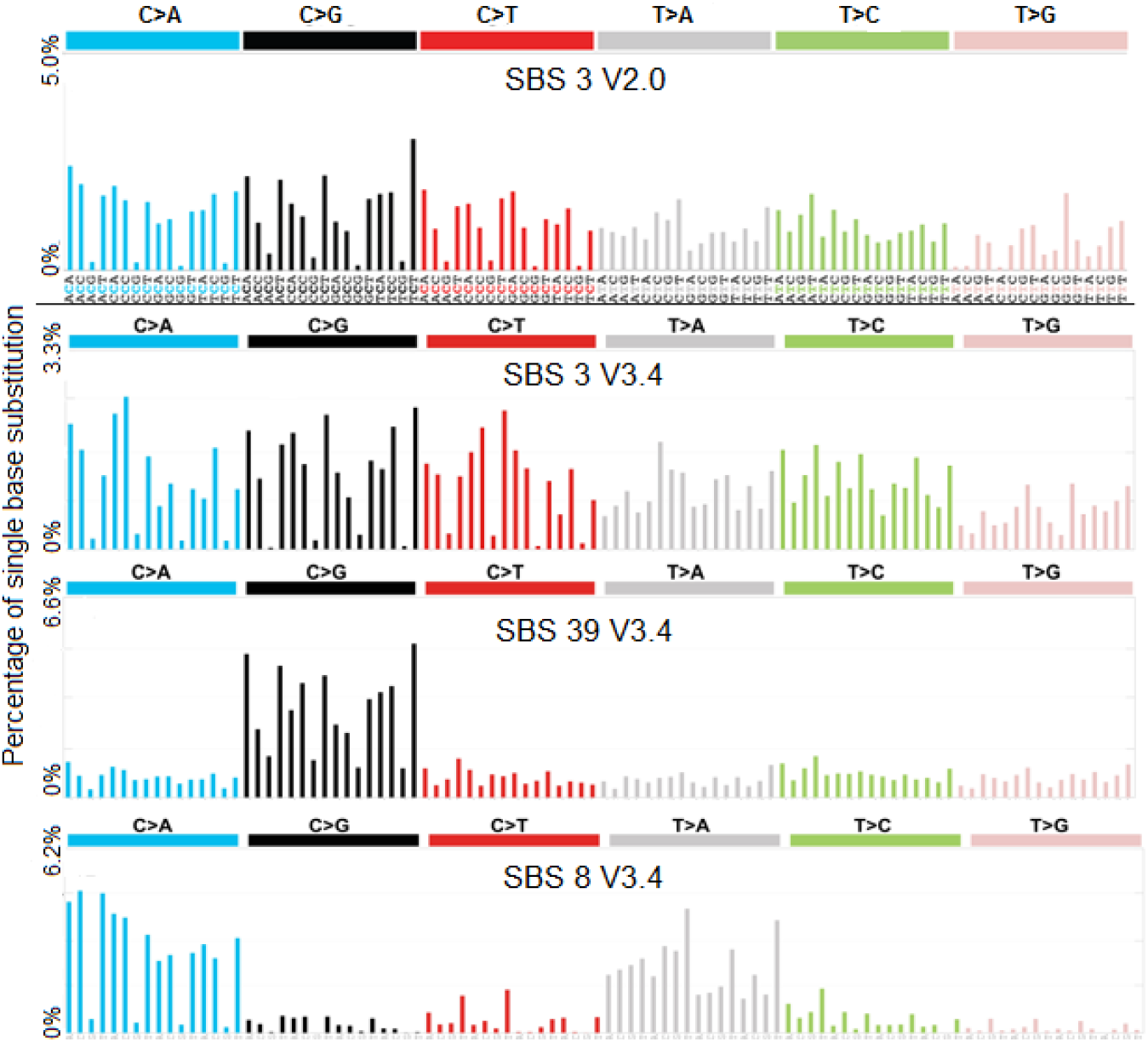
Mutational profile for SBS 3 V2.0, SBS 3 V3.4, SBS39 V3.4, and SBS 8 V3.4 using the conventional 96 mutation type classification. Each mutational profile is based on the six substitution types: C>A, C>G, C>T, T>A, T>C T>G, as well as the nucleotides immediately 5’ and 3’ to the mutation. Those plots were downloaded from the Cosmic mutational signature website (https://cancer.sanger.ac.uk/signatures/).

## Data Availability

All data produced in the present study are available upon reasonable request to the authors

## On-line Methods

### Participants, tissue samples, and sequencing data

Breast cancerparticipants were seen at City of Hope (COH), consented, and enrolled in an IRB-approved study. A total of 824 breast cancer cases were included in this study. All sequencing data were generated using the Ashion GEM ExTra assay (Exact Sciences). Ashion/Exact Sciences conducted the Whole-exome Sequencing with 400X average coverage for somatic variants using FFPE tumor samples (800X average coverage across 442 cancer genes and 300X over rest of genome) and 200X average coverage for germline variants using blood or skin tissue samples. All data are stored in the COH POSEIDON database. Tumor/germline sequence data were aligned to the human genome (Build37). Germline variant calling from the BAM files was performed using GATK Haplotype Caller. Somatic mutations were called using the FreeBays workflows[16] and further filtered by reads depth ≥ 10, and reads for alternative allele ≥ 4, and the alternate allele ratio ≥ 0.1. To reliably call single-base-pair (SBS) mutation signatures, samples with fewer than 10 somatic single nucleotide variants were excluded [12] leaving 763 samples for analyses.

### Pathogenic germline mutations and truncating somatic mutations in *BRCA1, BRCA2, PALB2*

#### Breast cancer participants

Germline variants in *BRCA1, BRCA2*, and *PALB2* were from clinical genetic testing of the Invitae Multi-Cancer panel. Forty-three of 763 patients carried pathogenic germline mutations including 14 in *BRCA1*, 19 in *BRCA2*, 1 in both *BRCA1* and *BRCA2*, and 9 in *PABL2*. An additional 15 patients had truncating somatic mutations (4 in *BRCA1*, 9 in *BRCA2*,1 in both *BRCA1* and *BRCA2*, and 1 in *PABL2*).

**Table 1.**
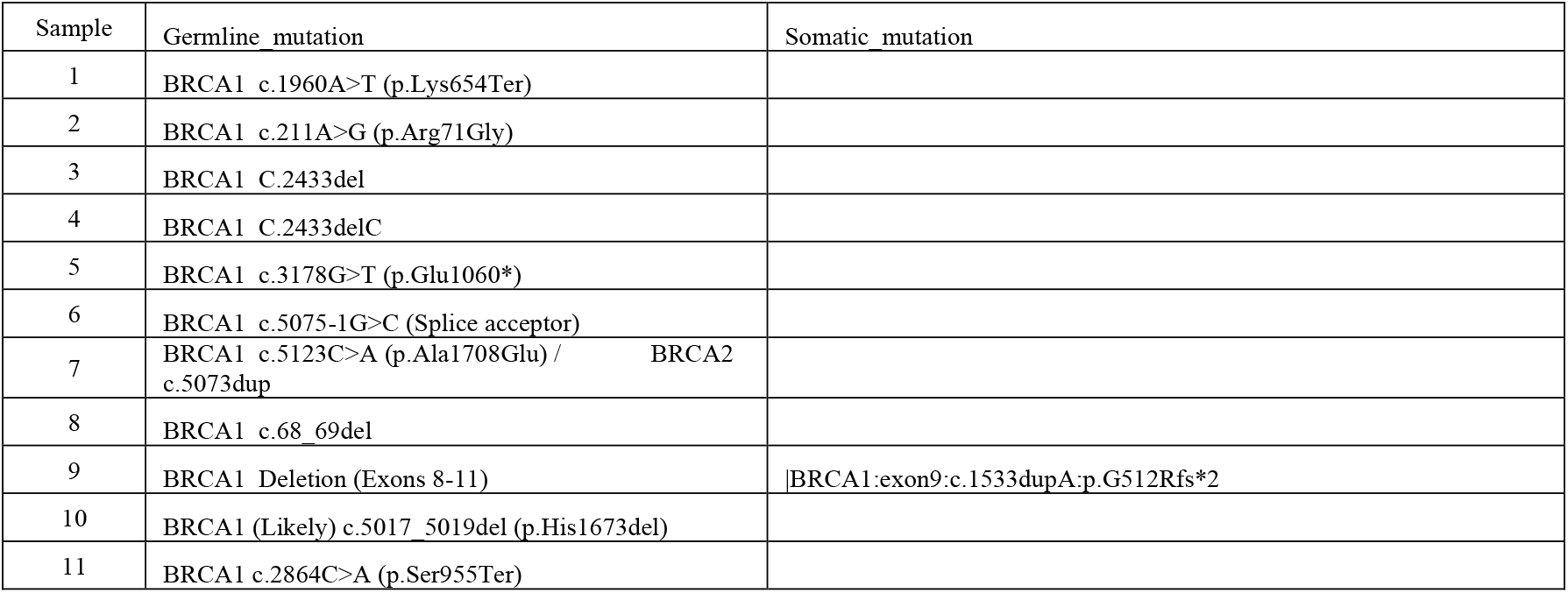
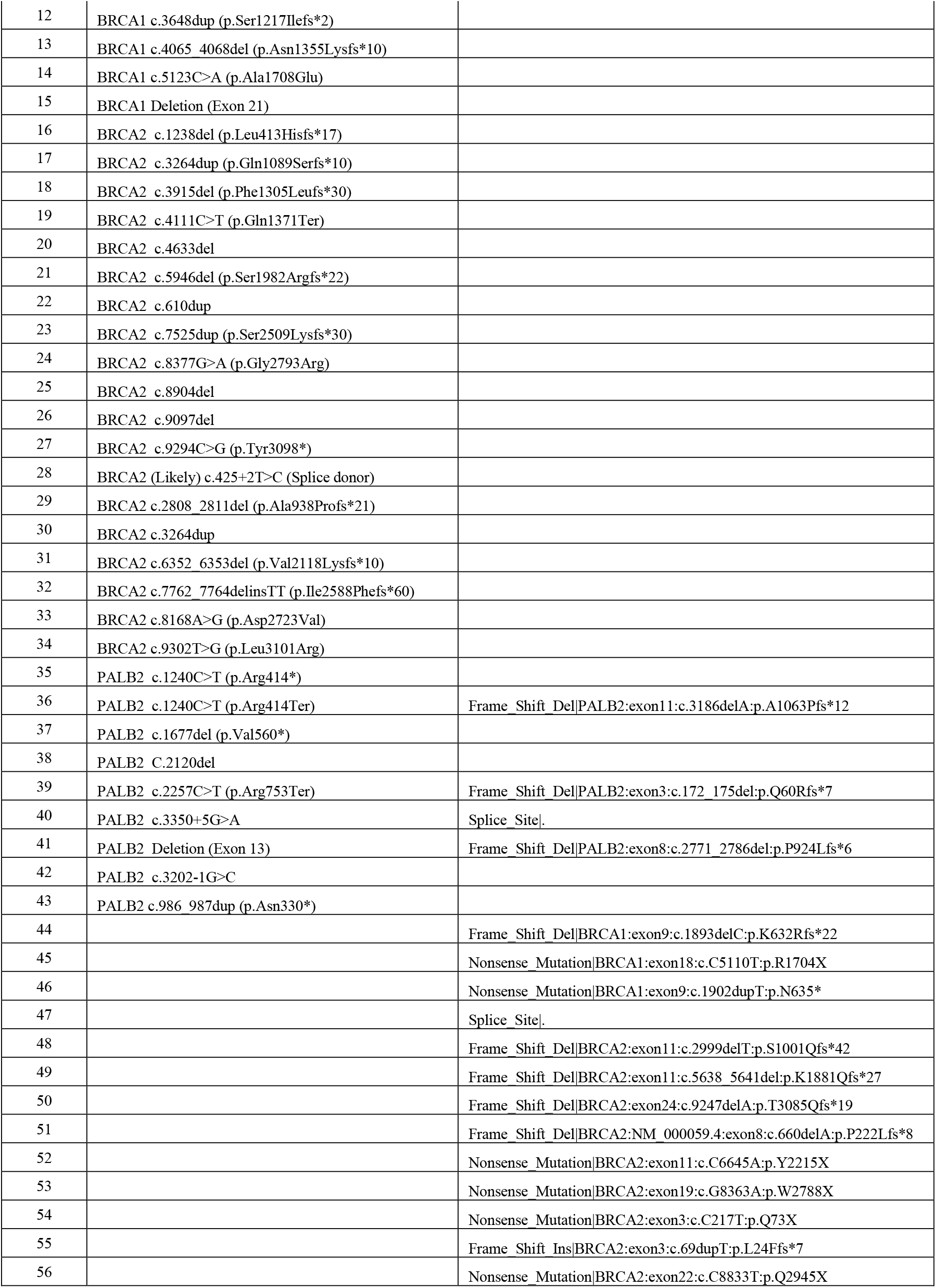
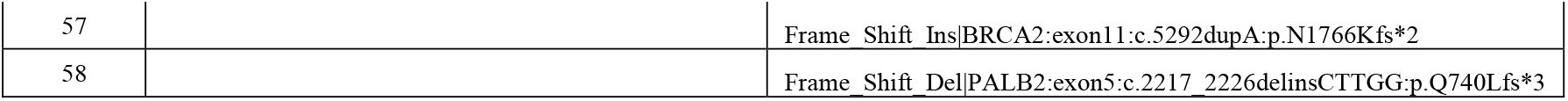
Pathogenic germline and truncating somatic mutations in 763 breast cancer samples from COH.

#### Ovarian cancer data

A mutation file for ovarian cancer data from TCGA project[7] was downloaded from the cbioportal (https://www.cbioportal.org/) database. Of the 313 samples, 50 samples had truncating germline mutations (25 in *BRCA1* and 25 in *BRCA2*) and 17 additional samples with truncating somatic mutations (9 in *BRCA1*, 6 in *BRCA2* and 2 in *PALB2*).

**Table 2.** Pathogenic germline mutation and truncating somatic mutations in 67 of 313 TCGA ovarian cancer samples from TCGA project.

| Tumor_Sample_Barcode | Mutation | Mutation Status |
| --- | --- | --- |
| TCGA-09-1669-01 | BRCA1_Frame_Shift_Del_p.E1346Kfs*20 | truncating germline mutation |
| TCGA-25-2392-01 | BRCA1_Frame_Shift_Del_p.E1346Kfs*20 | truncating germline mutation |
| TCGA-10-0931-01 | BRCA1_Frame_Shift_Del_p.E23Vfs*17 | truncating germline mutation |
| TCGA-13-1408-01 | BRCA1_Frame_Shift_Del_p.E23Vfs*17 | truncating germline mutation |
| TCGA-23-1027-01 | BRCA1_Frame_Shift_Del_p.E23Vfs*17 | truncating germline mutation |
| TCGA-23-1118-01 | BRCA1_Frame_Shift_Del_p.E23Vfs*17 | truncating germline mutation |
| TCGA-23-2078-01 | BRCA1_Frame_Shift_Del_p.E23Vfs*17 | truncating germline mutation |
| TCGA-23-2079-01 | BRCA1_Frame_Shift_Del_p.E23Vfs*17 | truncating germline mutation |
| TCGA-61-2109-01 | BRCA1_Frame_Shift_Del_p.K654Sfs*47 | truncating germline mutation |
| TCGA-04-1356-01 | BRCA1_Frame_Shift_Del_p.N723Ifs*13 | truncating germline mutation |
| TCGA-09-2045-01 | BRCA1_Frame_Shift_Del_p.Q1779Nfs*14 | truncating germline mutation |
| TCGA-57-1582-01 | BRCA1_Frame_Shift_Del_p.R1726Kfs*3 | truncating germline mutation |
| TCGA-13-0903-01 | BRCA1_Frame_Shift_Del_p.R504Vfs*28 | truncating germline mutation |
| TCGA-24-1470-01 | BRCA1_Frame_Shift_Del_p.T1677Ifs*2 | truncating germline mutation |
| TCGA-24-2298-01 | BRCA1_Frame_Shift_Ins_p.Q1395Lfs*11 | truncating germline mutation |
| TCGA-09-2051-01 | BRCA1_Frame_Shift_Ins_p.Q1756Pfs*74 | truncating germline mutation |
| TCGA-13-0883-01 | BRCA1_Frame_Shift_Ins_p.Q1756Pfs*74 | truncating germline mutation |
| TCGA-23-1122-01 | BRCA1_Frame_Shift_Ins_p.Q1756Pfs*74 | truncating germline mutation |
| TCGA-23-2077-01 | BRCA1_Frame_Shift_Ins_p.Q1756Pfs*74 | truncating germline mutation |
| TCGA-23-2081-01 | BRCA1_Frame_Shift_Ins_p.Q1756Pfs*74 | truncating germline mutation |
| TCGA-25-2401-01 | BRCA1_Frame_Shift_Ins_p.Q1756Pfs*74 | truncating germline mutation |
| TCGA-59-2348-01 | BRCA1_Nonsense_Mutation_p.E797* | truncating germline mutation |
| TCGA-13-0893-01 | BRCA1_Nonsense_Mutation_p.R504* | truncating germline mutation |
| TCGA-61-2008-01 | BRCA1_Nonsense_Mutation_p.W1815* | truncating germline mutation |
| TCGA-13-1494-01 | BRCA1_Splice_Site_p.X45_splice | truncating germline mutation |
| TCGA-13-0913-01 | BRCA2_Frame_Shift_Del_p.E1857Nfs*2 | truncating germline mutation |
| TCGA-13-0793-01 | BRCA2_Frame_Shift_Del_p.G3086Tfs*24 | truncating germline mutation |
| TCGA-25-2404-01 | BRCA2_Frame_Shift_Del_p.K343Nfs*6 | truncating germline mutation |
| TCGA-25-1318-01 | BRCA2_Frame_Shift_Del_p.L1491Qfs*12 | truncating germline mutation |
| TCGA-24-1417-01 | BRCA2_Frame_Shift_Del_p.N1706Lfs*5 | truncating germline mutation |
| TCGA-57-1584-01 | BRCA2_Frame_Shift_Del_p.N1784Hfs*2 | truncating germline mutation |
| TCGA-13-0900-01 | BRCA2_Frame_Shift_Del_p.N257Kfs*17 | truncating germline mutation |
| TCGA-13-0886-01 | BRCA2_Frame_Shift_Del_p.S1982Rfs*22 | truncating germline mutation |
| TCGA-13-1498-01 | BRCA2_Frame_Shift_Del_p.S1982Rfs*22 | truncating germline mutation |
| TCGA-13-1499-01 | BRCA2_Frame_Shift_Del_p.S1982Rfs*22 | truncating germline mutation |
| TCGA-24-2280-01 | BRCA2_Frame_Shift_Del_p.S1982Rfs*22 | truncating germline mutation |
| TCGA-59-2351-01 | BRCA2_Frame_Shift_Del_p.S1982Rfs*22 | truncating germline mutation |
| TCGA-04-1336-01 | BRCA2_Frame_Shift_Del_p.T1738Ifs*2 | truncating germline mutation |
| TCGA-24-2288-01 | BRCA2_Frame_Shift_Del_p.V220Ifs*4 | truncating germline mutation |
| TCGA-24-2024-01 | BRCA2_Frame_Shift_Del_p.Y1710* | truncating germline mutation |
| TCGA-24-1463-01 | BRCA2_Frame_Shift_Ins_p.I605Nfs*11 | truncating germline mutation |
| TCGA-04-1367-01 | BRCA2_Nonsense_Mutation_p.E294* | truncating germline mutation |
| TCGA-23-1026-01 | BRCA2_Nonsense_Mutation_p.K3326* | truncating germline mutation |
| TCGA-13-1512-01 | BRCA2_Nonsense_Mutation_p.K3326* | truncating germline mutation |
| TCGA-24-1562-01 | BRCA2_Nonsense_Mutation_p.K3326* | truncating germline mutation |
| TCGA-13-0726-01 | BRCA2_Nonsense_Mutation_p.R2394* | truncating germline mutation |
| TCGA-24-2293-01 | BRCA2_Nonsense_Mutation_p.R2520* | truncating germline mutation |
| TCGA-24-0975-01 | BRCA2_Splice_Site_p.X211_splice | truncating germline mutation |
| TCGA-24-1555-01 | BRCA2_Splice_Site_p.X2878_splice | truncating germline mutation |
| TCGA-25-1634-01 | BRCA2_Splice_Site_p.X2878_splice | truncating germline mutation |
| TCGA-25-1630-01 | BRCA1_Frame_Shift_Del_p.A521Qfs*11 | truncating somatic mutation |
| TCGA-24-2035-01 | BRCA1_Frame_Shift_Del_p.G1710Efs*4 | truncating somatic mutation |
| TCGA-13-1489-01 | BRCA1_Frame_Shift_Ins_p.N1265Kfs*4 | truncating somatic mutation |
| TCGA-25-1632-01 | BRCA1_Frame_Shift_Ins_p.S1217Rfs*21 | truncating somatic mutation |
| TCGA-25-1625-01 | BRCA1_Nonsense_Mutation_p.E116* | truncating somatic mutation |
| TCGA-29-2427-01 | BRCA1_Nonsense_Mutation_p.L431* | truncating somatic mutation |
| TCGA-04-1357-01 | BRCA1_Nonsense_Mutation_p.Q1538* | truncating somatic mutation |
| TCGA-13-0730-01 | BRCA1_Nonsense_Mutation_p.R1835* | truncating somatic mutation |
| TCGA-13-0761-01 | BRCA1_Splice_Site_p.X1495_splice | truncating somatic mutation |
| TCGA-13-0885-01 | BRCA2_Frame_Shift_Del_p.K1406Nfs*3 | truncating somatic mutation |
| TCGA-23-1120-01 | BRCA2_Frame_Shift_Del_p.P3278Lfs*35 | truncating somatic mutation |
| TCGA-13-0890-01 | BRCA2_Frame_Shift_Del_p.S1230Lfs*9 | truncating somatic mutation |
| TCGA-13-1481-01 | BRCA2_Frame_Shift_Del_p.S2697Kfs*31 | truncating somatic mutation |
| TCGA-04-1331-01 | BRCA2_Nonsense_Mutation_p.C711* | truncating somatic mutation |
| TCGA-09-2050-01 | BRCA2_Nonsense_Mutation_p.S1882* | truncating somatic mutation |
| TCGA-13-1477-01 | PALB2_Nonsense_Mutation_p.E263* | truncating somatic mutation |
| TCGA-09-2056-01 | PALB2_Nonsense_Mutation_p.Q370* | truncating somatic mutation |

#### Mutation signature assignment

We assigned SBS signatures for V2.0 and V3.4 using both the SigProfilerAssignment with high specificity and the MutationalPatterns with high sensitivity[10]. For signature assignment using SigProfilerAssignment tool, we directly submitted a matrix of somatic mutations for all 763 samples to the web tool, https://cancer.sanger.ac.uk/signatures/assignment/, developed by Diaz-Gay M. et al[10]. For signature assignment using MutationalPatterns, to avoid signature overfitting, we first identified a subset of signatures in each signature version specific to the somatic mutation data. Four and two signatures were extracted *de novo* from the breast and ovarian somatic mutation matrix data set, respectively, using the Non-Negative Matrix Factorization (NNMF) method[17] implemented in the MutationalPatterns R package[9]. Those signatures identified by NNMF were not novel because they showed high similarity to the known signatures in the Cosmic database. For example, the four signatures specific to the breast cancer somatic data showed high cosine similarity (> 0.6) with 15 signatures in version-2 Cosmic database and 25 signatures in version 3.4, respectively. Then we ran the signature refitting function in the MutationalPatterns R package to quantify the optimal contribution of the subset of signatures in each version (15 of 30 version-2.0 signatures and 25 of 86 version-3.4 signatures) to profile the mutation signatures of the 763 breast cancer samples. This signature refitting function assigned relative signature values for a set of Cosmic SBS signature to a sample by solving a non-negative least-squares constrains of the optimal linear combination of the set Cosmic signatures that most closely reconstructs the mutation profile matrix of the breast cancer sample[18].

### Selection of signatures relevant to *BRCA1*/*BRCA2*/*PALB2* mutations

We identified all signatures relevant to *BRCA1*/*BRCA2*/*PALB2* mutations instead of just selecting a minimal set of signatures to optimally predict *BRCA1*/*BRCA2*/*PALB2* mutation status. A random forest classification and feature selection algorithm, implemented in the Boruta R package[11], was used to select signatures relevant to *BRCA1*/*BRCA2*/*PALB2* mutations. In the selection process, mutation status (Yes or No) was the outcome variable, the set of signatures (for example, the 86 signatures in the version 3.4 assigned by the SigProfilerAssignment or the 25 of 86 version-3.4 signatures assigned by the MtuationalPatterns) were the list of explanatory variables. The maximum number of random forest run was set to 2000; The Boruta program iteratively discards a signature that is not relevant as assessed by a statistical test to be less important than its shadow signature created by randomly shuffling values of the original signature across samples[11]. Therefore, the shadow signatures created by permutation method is used to generate empirical null distribution of feature importance or relevance score. The relevance rank for all input signatures was determined by comparing the relevance of real signatures with the maximal, average and minimal relevance of shadow signatures (Supplementary Figure 1). if the relevance or importance of a signature could not be confirmed or rejected by 2000 iterations, the signature was considered as tentative important. Boruta was designed to find all relevant signatures, including weakly relevant signatures with importance value slightly greater than the maximal shadow relevance (see the “shadowMax” in the decision legend in the middle of two plots in the Supplementary Figure 1).

